# A Systematic Background Check of TRICARE Provider Names

**DOI:** 10.1101/2023.08.14.23294073

**Authors:** David Bychkov

## Abstract

During Covid-19, the Defense Health Agency’s TRICARE insurance plan expanded its coverage to include 30.1% additional civilian healthcare providers. The DHA’s Annual Report, however, states that TRICARE’s provider directories are only 80% accurate. Although the DHA’s 9.6 million beneficiaries need expanded access to care, they also require protection from misleading information, medical fraud, patient abuse, and identity theft. Since 2013, the Department of Health and Human Services’ Office of the Inspector General has excluded 17,706 physicians from federal health programs due to misconduct. Patients who receive care from excluded providers experience worse medical outcomes. To determine if any excluded provider names were found on TRICARE’s website, we performed background checks on TRICARE West’s healthcare provider directory between January 1 and March 2023. Out of 39,463 provider names sampled from 22 states, there were 2,398 matches (6.08%) with individuals and businesses found in the OIG List of Excluded Individuals and Entities (OIG-LEIE), the GSA-SAM, the HHS HIPAA Breach Report, the International Trade Administration’s Consolidated Screening List, the OIG-HHS Fugitive List, the FBI’s January 6th Capitol Violence List of Charged Defendants, State Medicaid Exclusion Lists, and FDA Debarment Lists. Our study includes demographic analysis of the matching names and recommendations for an Insider Threat Management model. We recommend that DHA officials publish the National Provider Identification (NPI) numbers of all TRICARE providers. NPI numbers facilitate more accurate background checks of healthcare providers.

## Introduction

In response to Covid-19, the Military Health System expanded access to telehealth and civilian providers. From 2020 to 2021, TRICARE, the Defense Health Agency’s insurance scheme, maintained a steady beneficiary pool of 9.6 million military beneficiaries. Meanwhile, its civilian roster ballooned from 548,297 to 713,395 providers – a 30.11% increase over 1 fiscal year (Defense Health Agency, 2022). Medical practices receive no mandatory training to mitigate cyber threats. Although the Health Insurance Portability and Accountability Act of 1996 (HIPAA), Medicare and TRICARE require healthcare providers to perform security risk assessments, 17% of respondents to the 2021 HIMSS Healthcare Cybersecurity Survey reported zero budget for risk assessments (HIMSS, 2022). In the same survey, 83% of respondents had experienced a cyber-attack. Due to budget or logistical concerns, 26% had reduced their overall cybersecurity budget (HIMSS, 2022). Unfortunately, military consumers are 76% more likely than other adults to experience medical benefit fraud and identity theft (Newman & Ritchie, 2020). According to a report by the Office of the Inspector General of the Department of Defense (DODIG), the Defense Health Agency (DHA)’s Program Integrity (PI) office suspended medical record audits related to improper payments to TRICARE West during the pandemic due to a lack of in-person investigators (DODIG, 2022). Despite receiving 600 lead requests, opening 110 new cases and managing 693 active cases in 2021, the DHA PI has sanctioned no new healthcare providers since August 2020 (DHA-Contract Resource Management, 2022) (Defense Health Agency/TRICARE, 2023). According to the DHA’s 2021 Report to Congress, only 80% of the provider directory information published by TRICARE’s managed care support contractors (MCSC) are accurate (Defense Health Agency, 2022). Due to call center problems and challenges with the Defense Enrollment Eligibility Reporting System (DEERS), the percentage of healthcare provider contracts in compliance with TRICARE fluctuated between 79.5% and 94.1% during the first 47 months of the current T2017 contract (Defense Health Agency, 2022). TRICARE West’s provider referral website ensures beneficiaries, however, that all healthcare providers “must meet TRICARE and HNFS stringent quality and credentialing requirements (Defense Health Agency, 2022).” As National Provider Identification (NPI) numbers are the sole unique identifier for licensed clinicians within the United States, however, it’s difficult to independently confirm TRICARE’s roster accuracy.

To demonstrate the limitations of provider due diligence available to TRICARE’s beneficiaries, we performed background checks on 39,463 names listed on TRICARE West’s public provider directory. Of those, 2,398 (6.08%) of the sampled provider names matched names in ten federal and state exclusion, sanction, violation, and criminal databases. Within this group, 10 TRICARE West provider names matched the names on the OIG-HHS list of fugitives wanted for health care fraud, abuse, or child support obligations and 54 names matched the Department of Justice’s list of January 6 Capitol Breach Defendants. Poor vetting practices not only harm national security but jeopardize the privacy of TRICARE beneficiary medical data and create unfair, regional disparities. For example, TRICARE patients in Utah, Minnesota, Kansas, Colorado, and Washington face a disproportionate risk of being referred to providers whose names appear on exclusion lists. Whereas TRICARE West beneficiaries who live in Honolulu, HI have access to 9 Military Treatment Facilities (MTF), patients in Amarillo, Texas and Bonneville County, ID would have to drive over 100 miles to find an MTF alternative to a provider associated with an exclusion. As millions of service members and veterans have security clearances, they face a heightened risk of extortion and identity theft. TRICARE, therefore, needs to ensure that only trusted providers access their beneficiaries’ protected health information (PHI). Nicholas et al performed a cross-sectional study of 8,204 Medicare beneficiaries who received care from excluded providers. It revealed that patients treated by fraudsters experience a 13-23% increased risk of mortality and 11-30% higher risk of hospitalization (Nicholas et al., 2019). We recommend that the Defense Health Agency mandate the disclosure of provider NPI numbers, revoked licenses and matches on exclusion databases to beneficiaries with active security clearances. As the pandemic is officially over, the DHA Program Integrity administrators need to deploy a formal Insider Threat Management (ITM) program that is scalable to MTFs, Health Net Federal Services Inc (TRICARE West), Humana Inc (TRICARE East) and small medical practices.

Continuous personnel screening is an effective mitigation practice against insider threats. The Centers for Medicare and Medicaid (CMS) already require healthcare organizations to screen their providers against two sources at regular intervals: the Office of Inspector General’s (OIG) List of Excluded Individuals/Entities (LEIE) and the General Services Administration’s SAM.gov exclusion list (GSA-SAM) (Kumaraswamy et al., 2022). Direct or indirect federal reimbursement for goods or services rendered by an excluded individual or entity is prohibited by the False Claims Act, FAR 9.404 “Exclusions in the System for Award Management’’ and the Civil Monetary Penalties (Acquisition.gov, 2023). This includes reimbursement for salaries, benefits, or items claimed/billed by licensed healthcare providers or administrative personnel. Healthcare organizations cannot purchase goods or services from excluded entities and vendors without jeopardizing their own Federal contracts (Centers for Medicare and Medicaid Services (CMS), 2021). SAM.gov includes several federal contracting databases such as USDA-FNS, TREAS-OFAC, the OPM (Office of Personnel Management) and more. Hospitals and insurance carriers must include SAM.gov datasets in their exclusion screening processes for employees and contractors (Patel & Sharma, 2022). Billing federal healthcare programs for services rendered by excluded providers can result in a minimum penalty of $10,000 per instance (OIG-HHS, 2013). To automate this process, McKesson, a revenue cycle management (RCM) and electronic health record (EHR) software provider has integrated exclusion monitoring tools into their products (McKesson, 2022). As TRICARE still accepts billing claims by fax and mail, fraudsters can still thwart automated exclusion screening processes (Defense Health Agency/TRICARE, 2022).

Approximately 18% of all service members receive security clearances (Government Accountability Office, 2018). They need to be able to discuss their medical conditions frankly with providers, without the fear of data breach or blackmail by an adversary (Department of Defense, 2011). Service members have a reasonable expectation that TRICARE-credentialed healthcare providers are not fugitives from justice, in violation of international sanctions, a threat to national security, or associated with a cyber breach. Multiple public databases exist to search names with respect to each of these issues, including the Department of Health Human Services’ Office of Civil Rights’ HIPAA Breach List Affecting 500 Individuals or More (Department of Health and Human Services, 2023), the HHS-Office of the Inspector General List of Fugitives (Department of Health and Human Services – OIG, 2023), the International Trade Administration’s Consolidated Screening List (International Trade Administration, 2023), the FBI’s List of January 6^th^ Capitol Breach Defendants (Federal Bureau of Investigation (FBI), 2023), State Medicaid Exclusion Lists (Medi-Cal, 2023), and the FDA’s various debarment lists (Food and Drug Administration, 2023). Although this study involved manual searches, Provider Trust and Pre-Employ enable organizations to cross-reference multiple databases – including the OIG-LEIE and GSA-SAM – for a fee (ProviderTrust, 2023). Unfortunately, no companies offer free, continuous provider background monitoring to service members, veterans, or their families.

As hackers seek insiders to help them target medical practices for malware exploits, the DHA needs to scrutinize each element of their provider roster. Our study identified 2,398 providers whose names are associated with an OIG-LEIE or SAM exclusion. Within this group, 203 names appeared in up to 3 additional types of databases. We recommend that the DHA mandate and monitor the findings of continuous background checks for all their healthcare providers.

Furthermore, the DHA needs to remove the names of excluded physicians from their provider directories and publish NPI numbers next to the names of all physicians on their sanctioned provider list. As there are real health risks associated with referring patients to excluded providers, TRICARE needs to increase their confidence in the accuracy of their provider websites from 80% to 100%. Finally, TRICARE beneficiaries need additional tools to recognize symptoms of a data breach and resources to prevent medical identity theft (Federal Trade Commission (FTC), 2023).

## Method

We mined the first and last names, specialty type, practice type, company names, and contact information of TRICARE-credentialed civilian providers for all 22 states from TRICARE West’s provider directory between January 1-31, 2023, at: https://www.tricare-west.com/content/hnfs/home/tw/bene/provider-directory.html

We then cross-referenced the provider first, last and corporate names against eight categories of exclusion, sanction and violation lists: the OIG-LEIE and SAM federal exclusion lists, the Department of Health and Human Services’ Office of Civil Rights HIPAA Breach Report Affecting 500 Individuals or More, 15 State Medicaid exclusion lists, the International Trade Administration’s Consolidated Screening List (a list of parties for which the United States Government maintains restrictions on exports, reexports, or transfers of items), three FDA debarment lists (drug imports, food imports and drug product), the FBI List of January 6 Federal Defendants, and the OIG-HHS List of Fugitives (wanted for health care fraud, abuse or child support obligations). Neither the Defense Health Agency nor TRICARE West provide the NPI numbers of healthcare providers on their websites. We limited our search to the 836 most populous zip codes (38 per state). Among these zip codes, TRICARE West listed no healthcare providers in 55 zip codes – primarily in Wyoming.

Our de-identified dataset can be found here:

Bychkov De-Identified TRICARE West Exclusions for Publication.xlsx

Our search yielded 39,463 provider names (5.53% of the total TRICARE nationwide civilian roster) to serve as the basis for our study. To ensure that our datasets were valid, we removed duplicate entries and manually confirmed each provider’s contact information. Only providers with an active website, office phone number, advertising campaign or Google business profile were included in the study.

## Institutional Review Board/Human Resource Protection

No confidential data was collected or used. This study was conducted with an exemption from the Human Resource Protection Program of Defense Acquisition University received on January 30, 2023.

## Results

We searched TRICARE West’s website for all providers who are currently accepting patients and listed in the most populous 38 zip codes of the 22 states they serve: Alaska, Arizona, California, Colorado, Hawaii, Idaho, Iowa (except the Rock Island Arsenal area), Kansas, Minnesota, Missouri (except the St. Louis area), Montana, Nebraska, Nevada, New Mexico, North Dakota, Oregon, South Dakota, Texas (Amarillo, Lubbock and El Paso areas only), Utah, Washington, and Wyoming.

Out of 39,463 names listed in the 836 zip codes we focused on, 2,398 (6.08%) shared the same first and last names as excluded parties found in 10 federal and state regulatory watchlists **(Tab.1)**. Among those 2,398 names, 2,197 appear on the OIG-LEIE and 2,311 appear on the GSA-SAM. Within this cohort, 2 appear on the HHS OCR HIPAA Breach Report, 69 appear on the International Trade Administration’s Consolidated Screening List, 10 appear on the OIG-HHS Fugitive List, 15 appear on 3 FDA debarment lists, 53 appear on the Department of Justice’s January 6^th^ Capitol Breach Defendant list, and 54 appear on 15 State Medicaid exclusion lists (AK: 0, CA: 38, HI: 0, ID: 0, IA: 1, KS: 0, MN: 0, MO: 0, MT: 0, ND: 0, NE: 0, NV: 1, TX: 20, WA: 0, WY: 0). One name matched a provider listed on the Defense Health Agency’s Sanctioned Provider List.

Our search matched 1,997 providers with 2 exclusion types, 230 providers with 1 type, 158 providers with 3 exclusion types, 12 providers with 4 exclusion types, and 1 provider with 5 exclusion types. Providers with 2 or more exclusions are likely to have their first, last and corporate names appear on the OIG-LEIE and GSA-SAM exclusion lists. All names that appear on the three FDA debarment lists, the HHS OCR HIPAA Breach Report, the HHS-OIG Fugitive List, and the ITA’s Consolidated Screening List also appear on the OIG-LEIE or the GSA-SAM. Most providers with names on the January 6^th^ Capitol Breach defendant list (50 out of 54) appear on the GSA-SAM. Unfortunately, the OIG-LEIE and GSA-SAM are not a panacea for fraud prevention. Both lists fail to show 1 provider name from the South Dakota State Medicaid exclusion list. Furthermore, we were only able to collect Medicaid exclusions data from 15 out of 22 states. The US Department of Health and Human Services does not disclose Medicaid-related exclusions to non-healthcare providers. Despite the limitations of study, it’s clear that TRICARE administrators need to continuously search for and screen their providers against multiple databases.

Our results include information about provider specialty **(Fig. 1)** and diploma type **(Tab. 1)**. The top specialty was Family Medicine (13.5%), followed by Nurse Practitioner (6.5%), Internal Medicine (4.8%), Optometrist (4.1%) and Pediatrics (3.9%). The provider diplomas most associated with exclusions were MD (1288), DO (199), NP (181), LCSW (107) and OD (99). Diploma types with the fewest exclusions include MN (1), RBT (1), LCP (1), PT (1) and CRNA (1). Our results also included information on provider gender. Males accounted for 59.42%, while females accounted for 40%. Two providers did not report their gender. Our findings are consistent with the literature. According to Chen et al’s cross-sectional study assessing all physician exclusions from 2007 to 2017, the number of physician exclusions grew by 20% to include nearly 0.3% of all US physicians (Chen et al., 2018). Exclusions were more common in the West and Southeast census regions and among male physicians. Older physicians with specialty training, specifically family medicine and osteopaths, were more likely to be excluded.

**Fig. 1.**
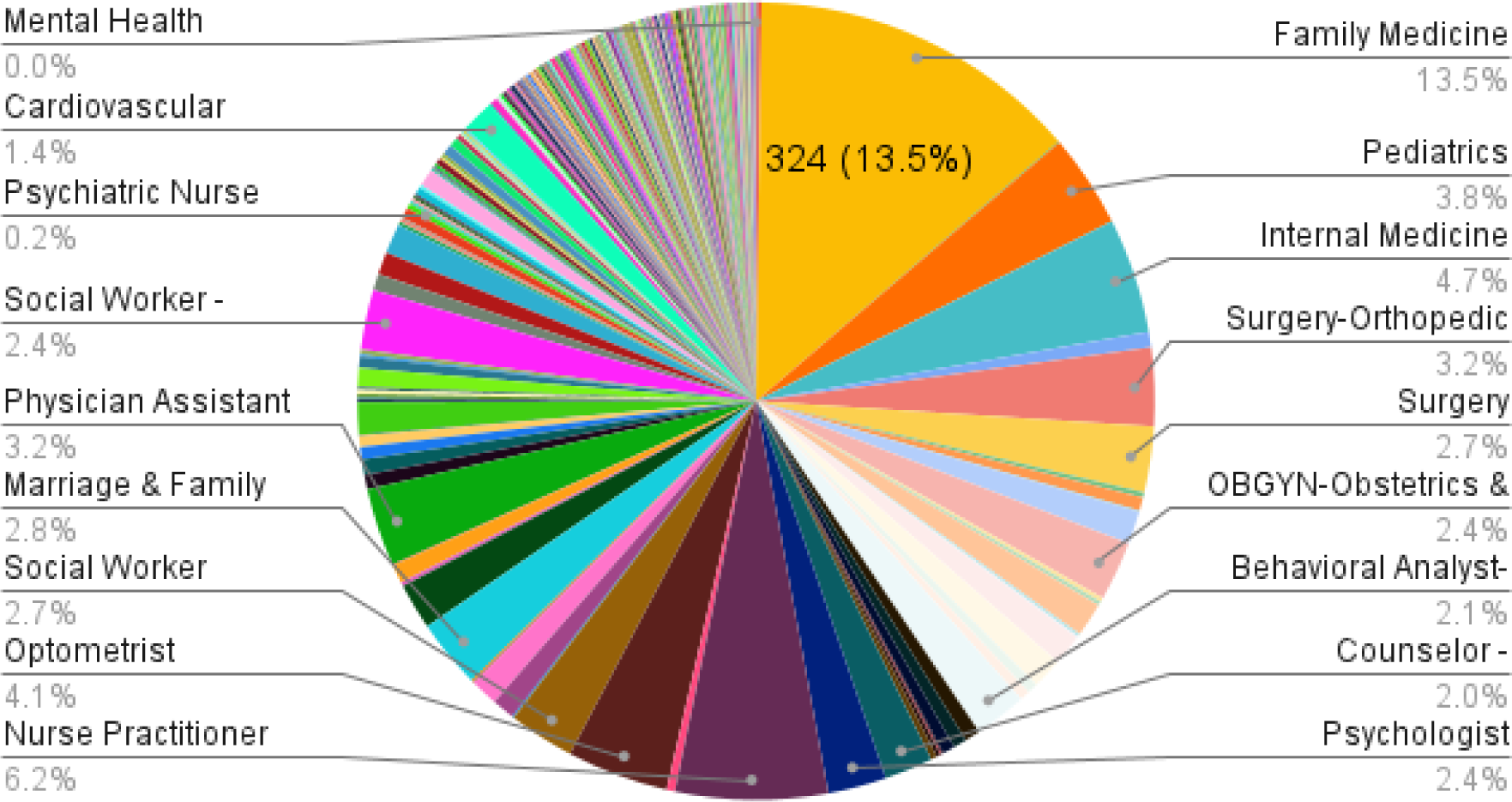
Matches by Specialty We conducted a search to identify the percentage of exclusions by specialty. Family Medicine was the top specialty (13.5%), followed by Nurse Practitioner (6.2%), Internal Medicine (4.7%), Optometrist (4.1%) and Pediatrics (3.8%).

**Tab. 1.** Top 5 Degrees of TRICARE West Providers Associated with Exclusions. We identified the top degree types associated with excluded providers. Accordingly, MD (1,288) was the most excluded provider type, followed by DO (199), NP (181), LCSW (107) and PA (99). Example diploma types with only one exclusion each include MN (1), RBT (1), LCP (1), PT (1) and CRNA (1).

| Top 5 Provider Degree | Total Number of Degree Holders |
| --- | --- |
| MD | 1,288 |
| DO | 199 |
| NP | 181 |
| LCSW | 107 |
| PA | 99 |

TRICARE West’s online referral tool includes the full addresses of each provider. We used this information to evaluate the impact of TRICARE West’s vetting practices on the local level. The states with the highest total number of providers associated with exclusions are UT (264), MN (227), KS (212), CO (201) and WA (180) **(Tab. 2)**. The states with the lowest total number of providers associated with exclusions are ID (32), MO (21), NE (15), MT (14) and WI (4). The Western states with the lowest number of TRICARE sanctioned providers since 1990 are: UT (0), MN, (0), CO (1), OR (0), MT (1), ND (0), SD (0) and WA (3). The most DHA sanctioned providers relevant to this study are in TX (16) and CA (11). According to the FY2021 Evaluation of the TRICARE Program, the average annual expenditure on a single beneficiary was $2,251 (Defense Health Agency, 2022). If the top 5% of our 2,398 identified providers by exclusion total were assigned 3 TRICARE beneficiaries each, the Defense Health Agency would have improperly paid out $810,360 to 120 physicians over the last calendar year. Under the False Claims Act, the agency could collect at least $10,000 per provider in penalties ($1,200,000) (US Department of Justice, 2023). By contrast, the 2022 DHA report on TRICARE shows only $900,000 in total collections from all excluded providers nationwide in calendar year 2020 and $100,000 in calendar year 2019 – down from a high of $1.4 million in 2017 (Defense Health Agency/TRICARE, 2023).

**Tab. 2.** State-by-State Multi-Database Search vs TRICARE Sanctioned Provider List. We studied exclusions in 22 states that offer TRICARE West. The states with the highest number of provider names associated with exclusions are UT (264), MN (227) and KS (212). Since 1990, the DHA has sanctioned 6 providers in Kansas, 1 in Colorado and 3 in Washington State. They sanctioned no providers in Utah or Minnesota.

| Top 5 States for Excluded Provider Matches | TRICARE Beneficiaries per State as per DHA 2021 Annual Report | TRICARE List of Sanctioned Providers (Offenses Originated with TRICARE) | Total TRICARE West Provider Names Matched Against 10 Exclusion Categories |
| --- | --- | --- | --- |
| Utah | 80,390 | 0 | 264 |
| Minnesota | 72,931 | 0 | 227 |
| Kansas | 120,503 | 6 | 212 |
| Colorado | 253,214 | 1 | 201 |
| Washington | 348,694 | 3 | 180 |
|  | Total: | 10 | Total: 1084 |

We analyzed 836 total zip codes within 22 states that offer TRICARE West **(Tab. 3)**. Five zip codes with the highest exclusions are 84096 – Herriman, UT (18), 84062 – Pleasant Grove, UT (17), 99669 – Soldotna, AK (16), 84790 – Washington County, Utah (14), and 80524 – Fort Collins, CO (14). Five zip codes with only 1 total exclusion are 96782 – Honolulu, HI (1), 79159 – Amarillo, TX (1), 84003 – Utah County, UT (1), 83401 – Bonneville County, ID (1) and 99505 – Anchorage, AK (1). We analyzed how many Military Treatment Facilities operate within a 100-mile radius of those zip codes with the highest concentration of excluded provider name matches. Whereas 96782 – Honolulu, HI has 9 MTFs within a 100-mile radius and 1 provider associated with an exclusion, 83401 – Bonneville County, ID, 84790 – Washington County, UT and 79159 – Amarillo, TX have no MTF alternatives to provider names associated with exclusions. In other words, TRICARE West beneficiaries in Honolulu face a much lower risk of seeing excluded providers. They can receive care directly at an MTF from DHA staff or service members who have cleared by the DoD’s Centralized Credentialing and Quality Assurance System (CCQAS), a Military Health System web application (Defense Health Agency/TRICARE, 2022).

**Tab. 3.** Top Zip Codes for Exclusions. We ranked 836 total zip codes within 22 states that offer TRICARE West based on the number of provider names associated with exclusions. We also indicate the number of Military Treatment Facilities within a 100-mile radius. The top 5 zip codes for exclusions are 84096 – Herriman, UT (18), 84062 – Pleasant Grove, UT (17), 99669 – Soldotna, AK (16), 66801 – Emporia, KS (14), and 80524 – Fort Collins, CO (14). For reference, we also show five zip codes with only 1 total exclusion. They include: 96782 – Honolulu, HI (1), 79159 – Amarillo, TX (1), 84003 – Utah County, UT (1), 83401 – Bonneville County, ID (1) and 99505 – Anchorage, AK (1). Whereas 96782 – Honolulu, HI has 9 MTFs and 1 provider associated with an exclusion within a 100-mile radius, 83401 – Bonneville County, ID and 79159 – Amarillo, TX have no MTF alternatives to the providers associated with exclusions.

| Top Zip Codes for Provider Exclusions | Total Provider Names Associated with Exclusions | Military Treatment Facilities within 100 Miles |
| --- | --- | --- |
| 84096 – Herriman, UT | 18 | 2 |
| 84062 – Pleasant Grove, UT | 17 | 2 |
| 99669 – Soldotna, AK | 16 | 2 |
| 84790 – Washington County, UT | 14 | 0 |
| 80524 – Fort Collins, CO | 14 | 2 |
| 96782 – Honolulu, HI | 1 | 9 |
| 79159 – Amarillo, TX | 1 | 0 |
| 84003 – Utah County, UT | 1 | 2 |
| 83401 – Bonneville County, ID | 1 | 0 |
| 99505 – Anchorage, AK | 1 | 2 |

## Discussion

Overall individual medical readiness among all non-deployed military components has dropped significantly since 2013. In Q4 2013, 75% of the Total Force reported being fully medically ready (e.g., completion of dental readiness assessments with satisfactory dental health, completion of periodic health assessments, deployment-limiting medical conditions status, current immunization status, completion of medical readiness lab tests, and possession of required individual medical equipment). That number dropped to 69% by Q4 FY2021. For defense leaders to rely on Total Force Medical Readiness reports, they need to trust the underlying methods of data collection. Based on our findings, hundreds of thousands of service members may have received health assessments, mental health reports, immunization reports, eye exams and lab tests during the pandemic from providers associated with exclusions. In future annual reports, the DHA needs to disclose how many beneficiaries received services from excluded providers. They also need to disclose how data collected from excluded providers are used in reports on military medical readiness.

TRICARE maintains a Sanctioned Provider List (Defense Health Agency/TRICARE, 2023). Whereas the OIG-LEIE contains 77,621 providers, the Defense Health Agency has sanctioned only 129 providers since 1990. In Oregon, Utah, Minnesota and the Dakotas, the DHA has sanctioned no providers. Meanwhile, our study of TRICARE West providers uncovered 2,398 possible leads that point to 4,697 federal and state combined exclusions, sanctions, and violations. As the TRICARE Sanctioned Provider List is an inadequate background check tool for beneficiaries, we recommend that the Defense Health Agency publish the NPI numbers of all civilian providers, require Health Net Federal Services (HNFS) and Humana to perform continuous background checks, and expand their list of sanctioned providers as appropriate.

TRICARE administrators and beneficiaries need reliable information to ensure patient privacy and safety. To help them avoid fraudsters, we also propose an Insider Threat Management Model **(Fig. 2)**. We recommend that TRICARE patients with Secret-level security clearance or higher never be referred to healthcare providers whose first, last and corporate name are associated with more than 1 exclusion in their state. If a search for “John + Smith” matches the name of an excluded provider on the OIG-LEIE and GSA-SAM in a beneficiary’s state, TRICARE administrators can reduce risk by recommending a different provider or suggesting the nearest MTF. If no nearby MTFs exist or if all available providers are associated with exclusions, TRICARE should pay for transportation to an MTF or treatment by a bona fide, out-of-network provider. According to our Insider Threat Management Model, any providers in the TRICARE network associated with 3 or more exclusions should have their beneficiaries transferred to an MTF. Patients who receive care from excluded providers face higher risks of mortality and hospitalization (Nicholas et al., 2019). Therefore, TRICARE needs to ensure beneficiaries with increased likelihood of being impacted are advised of their legal rights under HIPAA, including instructions on how to opt-out of the Joint Health Information Exchange, an electronic platform for transmitting TRICARE data to civilian providers (Defense Health Agency/TRICARE, 2020). Even if TRICARE eventually publishes provider NPI numbers, background checks are tedious and confusing to perform. Providers frequently change or add business names, last names, locations, and specialties. TRICARE.mil needs a comprehensive search tool that gathers data from all state, federal and licensing board databases, using a combination of spellings.

**Fig. 2.**
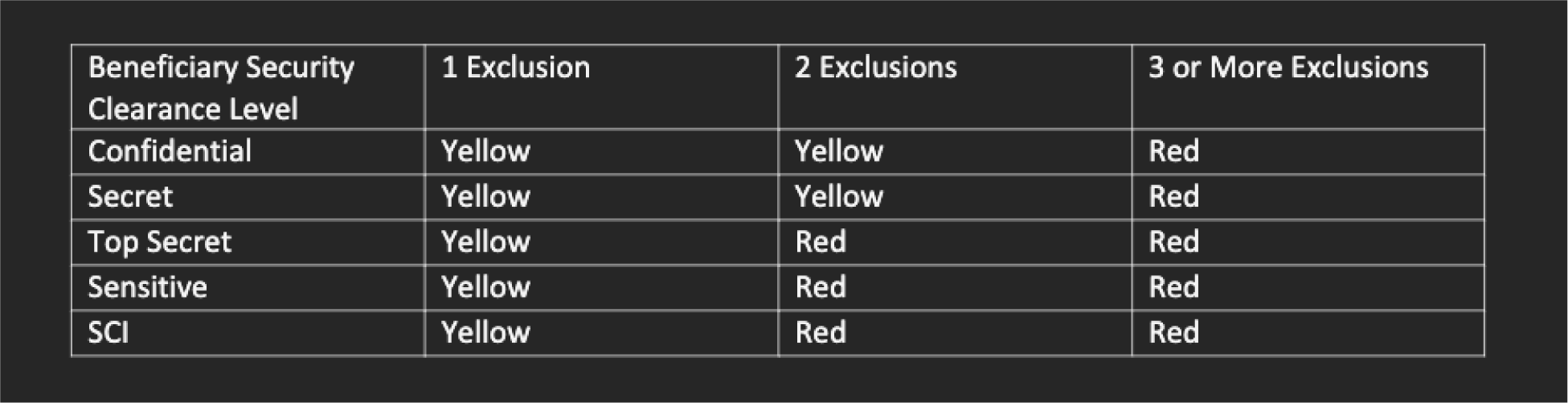
Insider Threat Management Model We propose an Insider Threat Matrix for TRICARE beneficiaries based on their security clearance level. Approximately 215,000 active-duty military officers have access to classified information and rely on TRICARE. TRICARE administrators need to ensure providers with exclusions have no access to these beneficiaries.

With a comprehensive, free search engine in place, TRICARE civilian providers can more easily comply with background check requirements. They can devote resources to third-party security risk assessments, protected health information (PHI) inventories, security information and event monitoring (SIEM) software, and HIPAA breach disclosures. By democratizing background check tools, the DHA can also generate new leads from future whistleblowers.

Patient outcomes data corroborate the need for restrictive background checks of new TRICARE providers. The Health Resources and Services Administration (HRSA) reported an increase in the number of adverse action reports and medical malpractice payments in several states within the TRICARE West region. In Utah, there were 359 adverse event reports in 2019 – the highest number recorded in the state’s history (Health Resources Services Administration National Practitioner Data Bank, 2023). In 2020, Wyoming experienced 137 adverse events – a 5-year record. In Montana, 67 providers were reported for adverse actions in 2021. By 2022, that number nearly doubled to 127. In 2013, there were 555 adverse events in Colorado – home of the DHA’s Program Integrity office. By 2020, adverse event reports peaked at 1,215. Adverse event trends recorded in HRSA’s National Practitioner Data Bank merit attention by regional fraud investigators and outreach campaigns to potential whistleblowers. With a 30.1% larger pool of subcontractors to oversee, the DHA needs clinician-specific, continuing medical education (CME) courses that convey the importance of information security, patient privacy, and medical misconduct. TRICARE should publicly recognize clinicians who undergo additional DHA training with virtual merit badges on their provider directory. Finally, DHA administrators can deliver these CMEs for free to their 713,395 civilian providers via Uniformed Services University for the Health Sciences (USUHS) and Defense Acquisition University (DAU).

## Limitations

Our study suffered from information and time constraints. The DHA states that only 80% of the public facing information of their provider directory is accurate (Defense Health Agency/TRICARE, 2023). We assume that the providers listed on TRICARE West’s roster as “accepting patients” were credentialed to render services to TRICARE patients and did so. In April 2023, the DHA announced they would replace HNFS with TriWest Healthcare Alliance as the prime contractor for TRICARE West. TriWest has made no announcements regarding the management of the TRICARE West provider referral website (Defense Health Agency, 2023). Currently, TRICARE West’s website provides no National Provider Identification (NPI) numbers for their clinicians. As it also fails to include their provider’s middle initials and a list of all states in which they are licensed, enormous effort was required to differentiate between common names. For example, our list of 2,398 matches includes a common female name that appears 10 times. Each of these females live in different zip codes and/or states, have different degrees, and practice completely unrelated types of medicine. In other words, hundreds of hours of manual, expert review was necessary to ensure our final sample was valid and free of duplicates.

To ensure our study hindered no ongoing law enforcement efforts, we disclosed all data and study materials to the Department of Defense Inspector General (DODIG) on May 8, 2023, and the DHA-OIG on May 24, 2023, in the form of a whistleblower complaint. In future studies, we intend to compare Medicare, TRICARE East and West, Children’s Health Insurance Program (CHIP) and SAMHSA provider rosters against additional exclusion, sanction, and violation categories (i.e., the Federal Sex Offender Registry). We also intend to develop products and interventions to automate background checks, protect patient privacy and educate healthcare administrators about insider threats.

## Conclusion

We compiled the full names, addresses, gender, degree, availability, and specialty types of 39,463 healthcare providers listed on TRICARE West’s public provider directory. Of those, 2,398 providers had first, last and/or corporate names that appeared in at least one of 10 types of federal or state exclusions databases. One of these names also appears on the DHA’s list of Sanctioned Providers. These 2,398 providers, representing 6.08% of TRICARE West’s total healthcare provider roster, practice primarily in Utah, Minnesota, Kansas, Colorado, and Washington. Under the False Claims Act and other laws, excluded providers are liable to repay the Defense Health Agency in addition to fines. To help TRICARE beneficiaries who face a disproportionate risk of being referred to an excluded provider, we recommend that the DHA re-impose site audits of medical practices, publish National Provider Identification (NPI) numbers, and utilize a formal Insider Threat Management model to protect patients with security clearances. TRICARE added hundreds of thousands of new civilian providers into the Military Health System to address Covid-19. They now need to provide mandatory training to these vendors. Newly credentialed TRICARE physicians need to become aware of their national security responsibilities. Furthermore, they need to be reminded about the patient harm caused by excluded, sanctioned, malicious and negligent providers. In summary, DHA administrators need to deliver accurate, reliable information to warfighters, veterans, and their families about their healthcare providers.

## Author notes

The views expressed in this article are those of the author. They do not reflect the official policy or position of the Department of Defense, the Defense Acquisition University or the U.S. Government. The author is currently a Department of Defense employee. This work was prepared as part of the author’s official duties as a Visiting Professor at Defense Acquisition University. Title 17 U.S.C. 105 provides that “Copyright protection under this title is not available for any work of the United States Government.” Title 17 U.S.C. 101 defines a U.S. Government work as a work prepared by a military service member or employee of the U.S. Government as part of that person’s official duties.

## Data Availability

All data produced in the present work are contained in the manuscript and available online at: https://docs.google.com/spreadsheets/d/1kzZzuKva_RaA8HftAPSkmaexxcef66XD/edit#gid=928032967

## Acknowledgments

The author would like to express gratitude to his former colleagues on the Cyber Enterprise Team at Defense Acquisition University. Dr. Paul Shaw provided substantial feedback and encouragement at each stage. He would also like to thank the men and women of the DODIG office for their willingness to review the data and re-review the data.

